# Effect of shared decision-making on trust in physicians in the management of systemic lupus erythematosus: The TRUMP2-SLE prospective cohort study

**DOI:** 10.1101/2024.01.05.24300886

**Authors:** Ryusuke Yoshimi, Nobuyuki Yajima, Chiharu Hidekawa, Natsuki Sakurai, Nao Oguro, Kenta Shidahara, Keigo Hayashi, Takanori Ichikawa, Dai Kishida, Yoshia Miyawaki, Ken-ei Sada, Yasuhiro Shimojima, Yuichi Ishikawa, Yuji Yoshioka, Yosuke Kunishita, Daiga Kishimoto, Kaoru Takase-Minegishi, Yohei Kirino, Shigeru Ohno, Noriaki Kurita, Hideaki Nakajima

**Author notes:** Noriaki Kurita and Hideaki Nakajima share last authorship. Correspondence to Ryusuke Yoshimi, MD, PhD, Department of Hematology and Clinical Immunology, Yokohama City University Graduate School of Medicine, 3-9 Fukuura, Kanazawa-ku, Yokohama 236-0004, Japan.

## Abstract

**Objectives:** Few studies have explored whether the involvement of patients in shared decision-making (SDM) is beneficial to the management of systemic lupus erythematosus (SLE). Therefore, this study investigated the relationship between patient participation in SDM and their trust in physicians using data from the TRUMP2-SLE study.

**Methods:** Data regarding the nine-item Shared Decision-Making Questionnaire (SDM-Q-9 scores), Trust in Physician Scale (TIPS) scores, and Abbreviated Wake Forest Physician Trust Scale (A-WFPTS) scores for interpersonal trust in a physician and trust in the medical profession were collected from patients with SLE who visited the outpatient clinics of five facilities in Japan through a self-administered questionnaire. The relationships between these scores were analyzed.

**Results:** This study included 433 patients with SLE. The median baseline TIPS and A-WFPTS (attending physician version) scores were 82 (73–93) and 80 (70–95), respectively. A higher baseline SDM-Q-9 score was correlated with an increase in the TIPS score at 1 year (adjusted mean difference per 10-pt increase, 0.95 pt [95%CI 0.18–1.71]). A higher baseline SDM-Q-9 score was also correlated with a higher A-WFPTS score for interpersonal trust (adjusted mean difference per 10-pt increase, 2.16 pt [1.41-2.92]). The baseline A-WFPTS (general physician version) score of 65 (50–80) was also correlated with an increase in the A-WFPTS score at 1 year (adjusted mean difference per 10-pt increase, 1.28 pt [0.43–2.14]).

**Conclusions:** Engagement of patients with SLE in SDM elevates their trust in the attending physicians and healthcare providers, potentially enhancing doctor-patient relationships and overall healthcare trust.

**KEY MESSAGES:** *What is already known about this subject?:*

➢ The importance of the involvement of patients in shared decision-making (SDM) in the management of systemic lupus erythematosus (SLE) has been emphasized as one of the principles in the EULAR recommendation.

*What does this study add?:*

➢ The baseline SDM-Q-9 scores were correlated with the Trust in Physician Scale (TIPS) and the attending physician version of the Abbreviated Wake Forest Physician Trust Scale (A-WFPTS) scores at 1 year.
➢ The baseline SDM-Q-9 scores at baseline were correlated with the general physician version of the A-WFPTS score at 1 year.
➢ These results indicate that the involvement of patients with SLE in SDM increases their trust in the attending physicians and physicians in general.

*How might this impact on clinical practice or future developments?:*

➢ This study demonstrated the importance of SDM in maintaining long-term physician-patient relationship during the management of SLE.
➢ Involvement of patients with SLE in SDM may improve the doctor-patient relationship and trust in healthcare.

## INTRODUCTION

Reflecting the growing recognition of the concept of shared decision-making (SDM) between patients and healthcare professionals in a variety of diseases, including rheumatic diseases [1], SDM in systemic lupus erythematosus (SLE) has been highlighted as one of the overarching principles in the recommendations and guidelines of the European Alliance of Associations for Rheumatology (EULAR) since 2019. [2,3] SDM is expected to aid patients with SLE whose treatment regimen requires modifications during each period of exacerbation and remission to achieve treatment goals. [4] However, few empirical studies have examined the pathway to the achievement of treatment goals. For instance, it is generally still largely unexplored whether an increase in patient satisfaction due to high-quality SDM ultimately leads to increased trust in physicians and adherence to physicians’ recommendations, [5] and this is particularly the case in patients with SLE.

Patient satisfaction and trust in physicians are expected outcomes of SDM in rheumatology. [5,6] Trust in physicians is central to medical care, as it aids in maintaining medication adherence among patients with SLE. [7–10] Good SDM is associated with trust in the physician among patients with lupus nephritis. [10] However, this association may have been confounded by education and disease activity. Moreover, the temporal relationship remains unknown owing to the cross-sectional design of the study, and it is unclear whether interpersonal physician-patient interactions affect patients’ trust in physicians in general. Thus, it is necessary to verify the effect of physicians’ efforts to promote SDM on patients’ trust in physicians, both interpersonally and generally, [11] via a well-designed study. [5]

Therefore, the present study evaluated the longitudinal association between the degree of patient participation in SDM and trust in physicians among patients with SLE in Japan using data from the Trust Measurement for Physicians and Patients with SLE (TRUMP2-SLE) project.

## METHODS

### Study design and setting

This prospective cohort study was conducted using data from the TRUMP2-SLE Project, a multipurpose cohort study conducted across five academic medical centers (Showa University Hospital, Okayama University Hospital, Shinshu University Hospital, Yokohama City University Hospital, and Yokohama City University Medical Center). This study adhered to the tenets of the Declaration of Helsinki and ethical guidelines for epidemiologic research in Japan and was approved by the Institutional Review Board of Yokohama City University (F220600012). All patients provided written informed consent to participate in the registry and for the publication of their data.

### Patients

The inclusion criteria were as follows: (i) patients with SLE aged ≥20 years who satisfied the 1997 ACR revised classification criteria; (ii) receiving rheumatology care at the participating center; and (iii) ability to respond to the questionnaire survey. All the patients and attending rheumatologists were Japanese. Patients with dementia or total blindness were excluded. Data extracted from the electronic medical records and self-administered questionnaires completed by the registered patients and their attending physicians between June 2020 and August 2021 was used in this study. All data were collected at the time of registration.

### Exposure

The primary exposure was SDM, which was measured using the nine-item Japanese version of the Shared Decision-Making Questionnaire (SDM-Q-9). [12,13] SDM-Q-9 measures the extent of the involvement of the patients in the decision-making process and is expected to be used in rheumatology care. [4,14] SDM-Q-9 comprises nine items scored on a six-point scale.

The patients selected one of the following responses for each item: ‘completely disagree’ (0 points) to ‘completely agree’ (5 points). The sum of the scores was converted to a score ranging from 0 to 100. The Cronbach’s alpha coefficient for the Japanese version of SDM-Q-9 was 0.917, and the construct validity of the questionnaire has been established. [12]

### Outcomes

The outcome measure was trust in physicians. The primary outcome measure was trust in the attending rheumatologist, which was measured using the Japanese version of the modified Trust in Physician Scale (TIPS). [15,16] This 11-item scale measures trust in the physician in terms of physicians’ dependability, trust in physicians’ knowledge and skills, and trust in confidentiality and reliability of the information exchanged between the physician and patient. Each item was scored on a five-point Likert scale. The patients selected one of the following responses for each item: ‘Totally disagree’ (1 point) to ‘Totally agree’ (5 points). The score for negatively worded items was inverted, and the sum of all scores was converted to a score ranging from 0 to 100. The Cronbach’s alpha coefficient for the scale was 0.91, and the construct validity of the scale has been established. [16]

The secondary outcome measures included trust in the attending physician and trust in physicians in general. These outcome measures were evaluated using the Japanese version of the Abbreviated Wake Forest Physician Trust Scale (A-WFPTS). [17] A-WFPTS measures competence, honesty, fidelity, and general trust on a five-point Likert scale. [18] The patients selected one of the following responses for each item: ‘Strongly disagree’ (1 point) to ‘Strongly agree’ (5 points). The scores for the negatively worded item were inverted, and the sum of all item scores was converted to a score ranging from 0 to 100. The Cronbach alpha coefficients for the A-WFPTS for trust in the attending physician and trust in physicians in general were 0.85 and 0.88, respectively, and the construct validity of these scales has been established. [17]

### Measurement of covariates

Confounding variables included the variables suspected to affect SDM and trust in physicians based on evidence in the literature and expert medical knowledge. Age, [17] sex, [19] marital status, final education, [17] household income, [16] disease activity, [19] duration of illness [20], and the period in charge of the attending physician were the included variables. The Systemic Lupus Erythematosus Disease Activity Index 2000 (SLEDAI-2K) was used by the attending physician to measure disease activity. Each trust level in the physicians’ scale at baseline was also included as a confounding variable.

### Statistical analysis

All statistical analyses were performed using Stata/SE, version 16.1 (StataCorp, College Station, TX, USA) and IBM SPSS Statistics for Windows version 29.0 (IBM Corp., Armonk, NY, USA). Summary statistics were used to describe demographic, clinical, and laboratory characteristics. Normally distributed continuous variables are presented as mean ± standard deviation (SD), whereas non-normally distributed continuous variables are presented as median and interquartile range (IQR). Categorical and ordinal data are presented as proportions. A series of general linear models was fitted with the outcome variables as dependent variables and SDM-Q-9 as a continuous variable, adjusting for the aforementioned confounding variables. Multiple imputations with chained equations were performed 100 times to impute the missing values. [21]

## RESULTS

### Flow of the study

Among the 433 patients with SLE registered in the TRUMP2-SLE project (Figure 1), data regarding the TIPS, A-WFPTS (attending physician version), and A-WFPTS (general physician version) scores at 1 year were available for 413, 421, and 420 patients, respectively.

**Figure 1.**
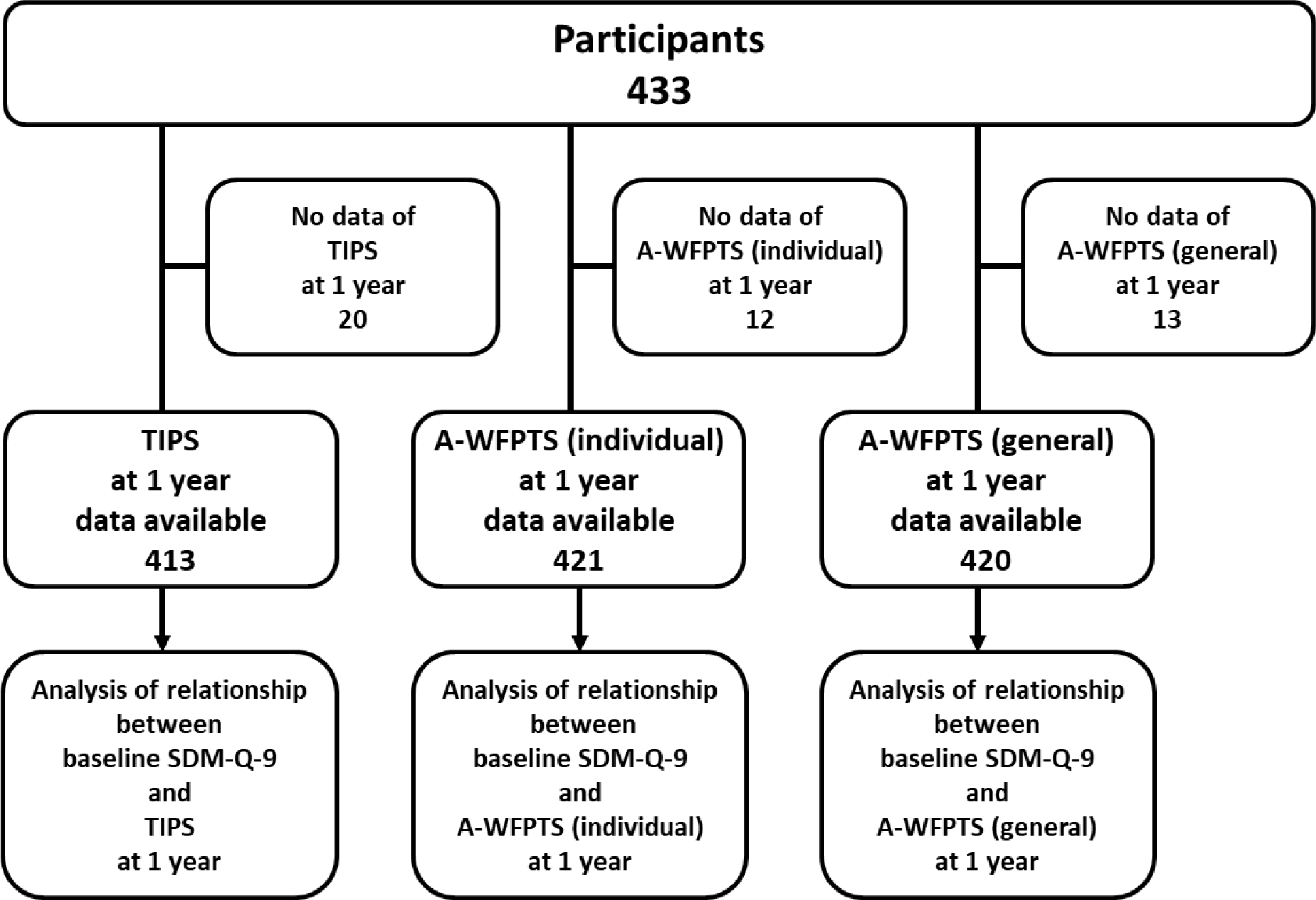
Patient enrollment. A total of 433 patients were included in the study. The associations between baseline SDM measures and trust in physicians at one year were analyzed in prospective cohort studies.

### Patient characteristics

The mean age of the enrolled patients was 46.2 ± 14.3 years, and 378 (87%) patients were women (Table 1). The median (IQR) duration of disease was 12.6 (6.5–19.9) years. More than half of the patients had been followed by the attending physicians for ≥3 years. The median (IQR) SLEDAI-2K score at the time of registration was 4 (range: 1–6). Most patients had an annual income of 5–10 million yen (39%), followed by 2.5–5 million yen (31%). Approximately 69% of patients had high school-, technical college-, or junior college-level education, whereas 26% of patients had university- or graduate school-level education. Marital status was reported for 54.7% of the participants. The median (IQR) SDM-Q-9 score was 78 (60–91).

**Table 1.**
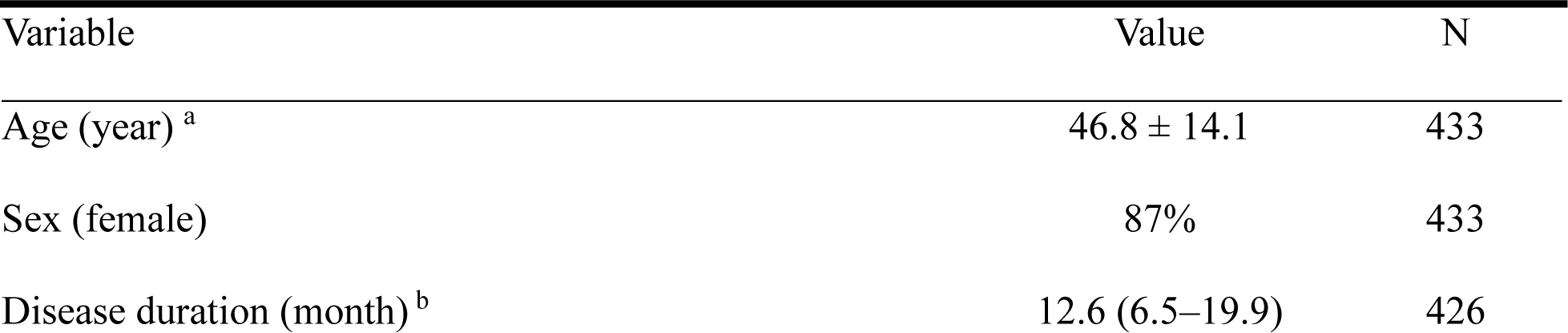
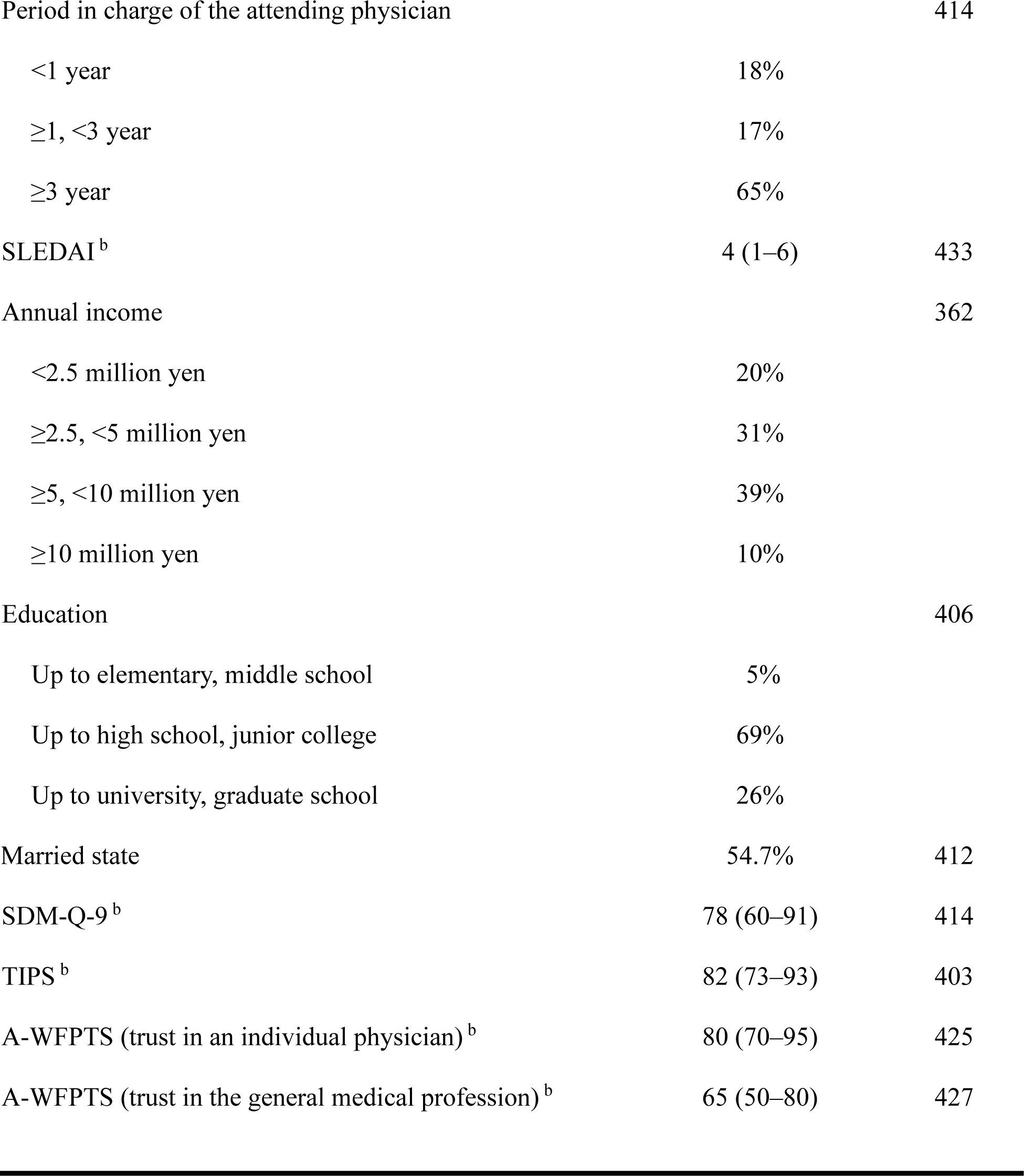
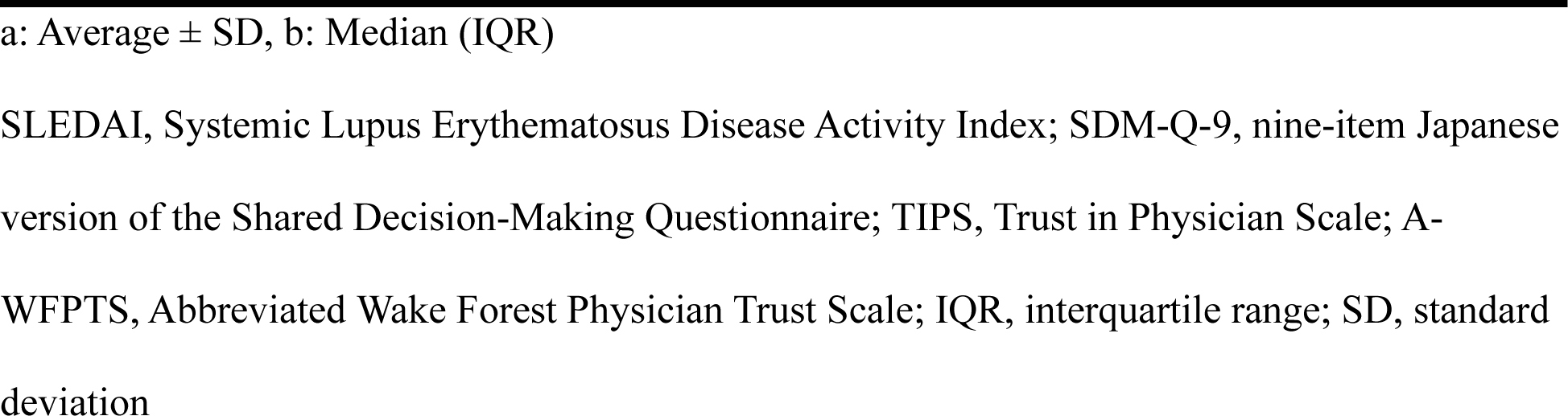
Patient characteristics.

### Association between SDM and trust in the rheumatologist at 1 year

The median (IQR) baseline TIPS and A-WFPTS (attending physician version) scores were 82 (73–93) and 80 (70–95), respectively. After adjustment, a greater baseline SDM-Q-9 score was associated with a greater TIPS score at 1 year (adjusted mean difference per 10-pt increase, 0.95 pt [95%CI 0.18–1.71]) (Table 2). Similarly, a greater baseline SDM-Q-9 score was associated with a greater A-WFPTS score for interpersonal trust in a physician at 1 year (adjusted mean difference per 10-pt increase, 2.16 pt [95%CI 1.41–2.92]) (Table 3).

**Table 2.** Baseline SDM-Q-9 score as an explanatory variable for TIPS at 1 year.

| Variable | Adjusted mean difference (95% CI) | P value |
| --- | --- | --- |
| SDM-Q-9 | 0.095 (0.018, 0.171) | 0.016* |
| Age (year) | 0.015 (-0.085, 0.114) | 0.771 |
| Sex (female) | 0.219 (-3.222, 3.660) | 0.900 |
| Disease duration (year) | 0.014 (-0.112, 0.139) | 0.831 |
| Period in charge of the attending physician<br>(vs. <1 year) | Ref. |  |
| ≥1, <3 year | -0.050 (-3.933, 3.834) | 0.980 |
| ≥3 year | -0.430 (-3.577, 2.717) | 0.788 |
| SLEDAI | -0.165 (-0.468, 0.138) | 0.285 |
| Annual income (vs. <2.5 million yen) | Ref. |  |
| ≥2.5, <5 million yen | -0.703 (-4.367, 2.960) | 0.706 |
| ≥5, <10 million yen | -1.311 (-5.134, 2.512) | 0.500 |
| ≥10 million yen | 1.626 (-3.600, 6.853) | 0.541 |
| Education (vs. up to elementary, middle<br>school) | Ref. |  |
| Up to high school, junior college | -1.229 (-7.541, 5.082) | 0.702 |
| Up to university, graduate school | -1.250 (-7.880, 5.379) | 0.711 |
| Marital status | -0.320 (-3.078, 2.437) | 0.819 |
| TIPS at baseline | 0.603 (0.486, 0.720) | <0.001*** |

\* $p < 0.05$ , \*\* $p < 0.01$ , \*\*\* $p < 0.001$ .
SLEDAI, Systemic Lupus Erythematosus Disease Activity Index; SDM-Q-9, nine-item Japanese version of the Shared Decision-Making Questionnaire; A-WFPTS, Abbreviated Wake Forest Physician Trust Scale; CI, confidence interval

**Table 3.**
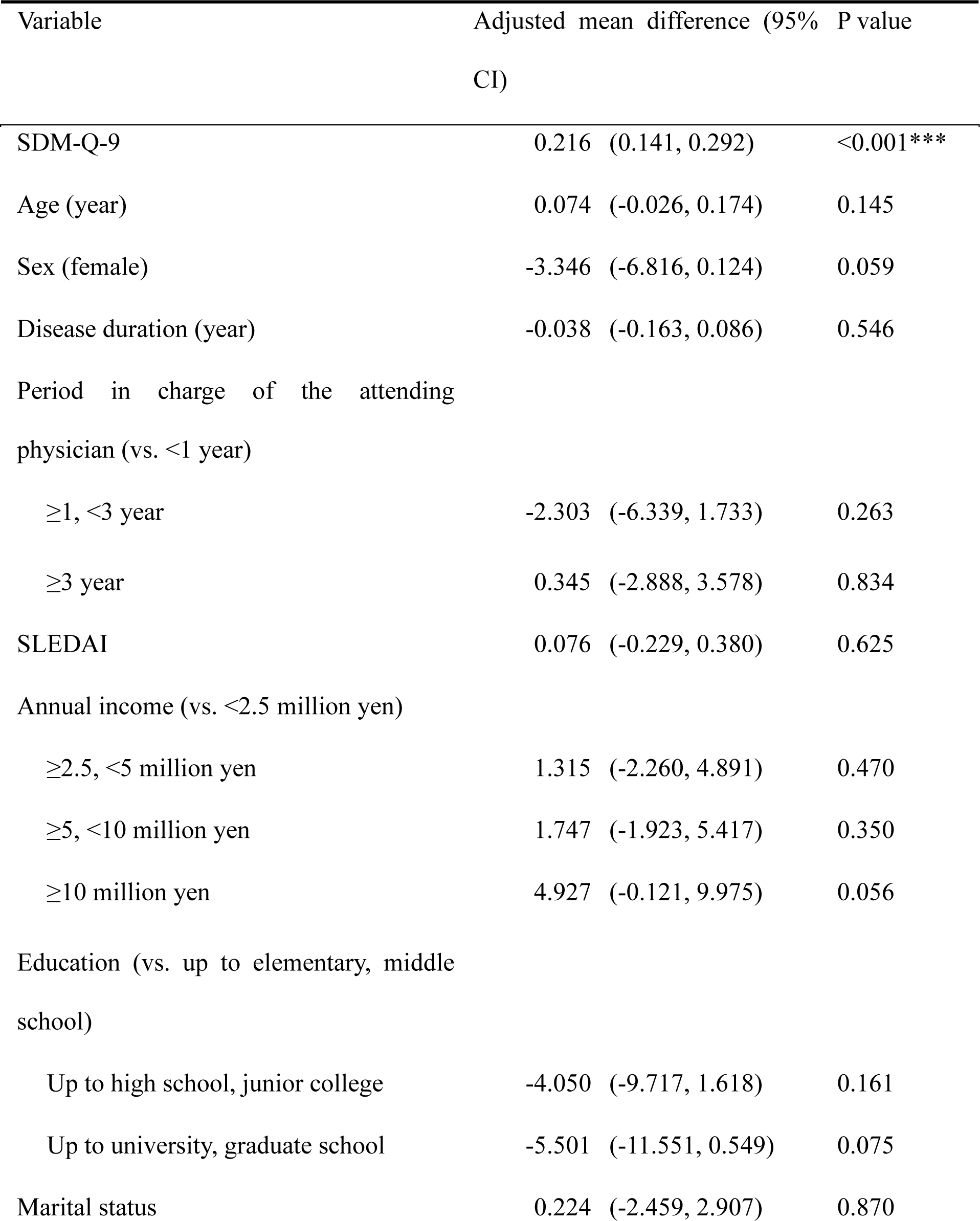
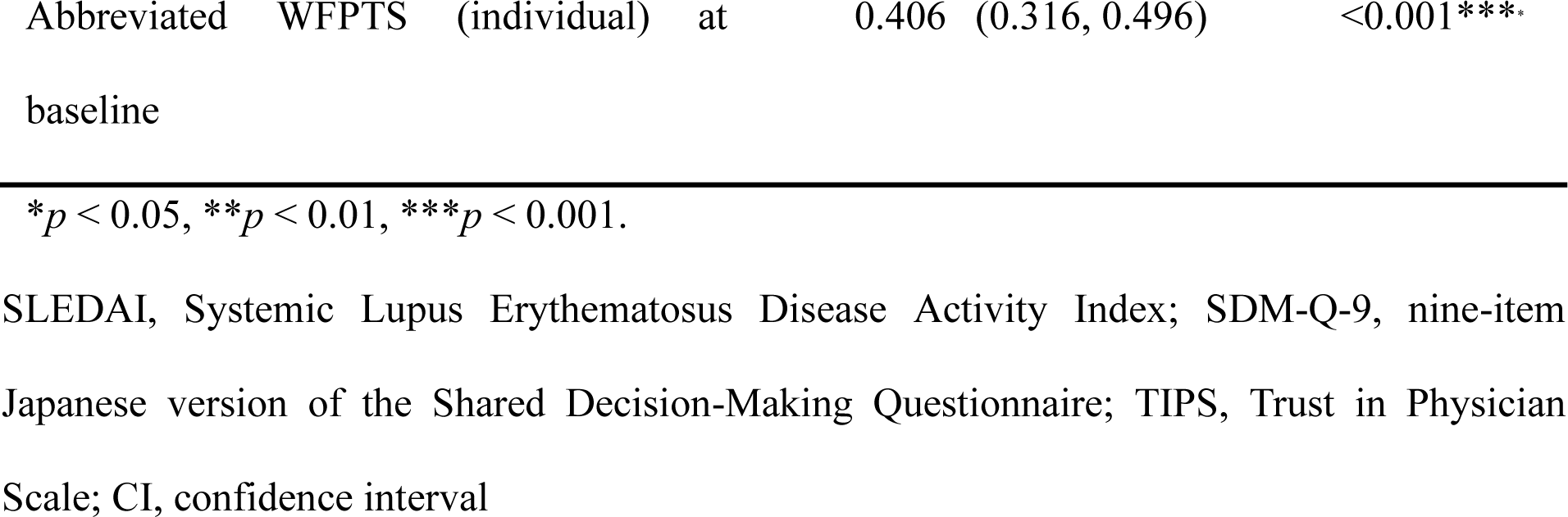
Baseline SDM-Q-9 score as an explanatory variable for abbreviated WFPTS (individual) at 1 year.

### Association between SDM and trust in general physician at 1 year

The median (IQR) baseline A-WFPTS (general physician version) score was 65 (50–80). After adjustment, a greater baseline SDM-Q-9 score was associated with a greater A-WFPTS score for trust in general physician at 1 year (adjusted mean difference per 10-pt increase, 1.28 pt [95%CI 0.43–2.14]) (Table 4).

**Table 4.** Baseline SDM-Q-9 score as an explanatory variable for abbreviated WFPTS (general) at 1 year.

| Variable | Adjusted mean difference (95% CI) | P value |
| --- | --- | --- |
| SDM-Q-9 | 0.128 (0.043, 0.214) | 0.003** |
| Age (year) | 0.203 (-0.069, 0.337) | 0.003** |
| Sex (female) | 1.926 (-2.620, 6.473) | 0.405 |
| Disease duration (year) | -0.076 (-0.238, 0.085) | 0.351 |
| Period in charge of the attending physician<br>(vs. <1 year) |  |  |
| ≥1, <3 year | -0.208 (-5.435, 5.020) | 0.938 |
| ≥3 year | -1.866 (-6.052, 2.320) | 0.381 |
| SLEDAI | -0.178 (-0.574, 0.218) | 0.377 |
| Annual income (vs. <2.5 million yen) |  |  |
| ≥2.5, <5 million yen | 0.324 (-4.468, 5.115) | 0.894 |
| ≥5, <10 million yen | -4.308 (-9.178, 0.563) | 0.083 |
| ≥10 million yen | -0.635 (-7.282, 6.013) | 0.851 |
| Education (vs. up to elementary, middle<br>school) |  |  |
| Up to high school, junior college | -0.560 (-7.952, 6.833) | 0.882 |
| Up to university, graduate school | -3.554 (-11.474, 4.366) | 0.378 |
| Marital status | 0.569 (-2.997, 4.135) | 0.754 |
| Abbreviated WFPTS (general) at baseline | 0.467 (0.387, 0.547) | <0.001*** |

\* $p < 0.05$ , \*\* $p < 0.01$ , \*\*\* $p < 0.001$ .
SLEDAI, Systemic Lupus Erythematosus Disease Activity Index; SDM-Q-9, nine-item Japanese version of the Shared Decision-Making Questionnaire; TIPS, Trust in Physician Scale; CI, confidence interval

## DISCUSSION

The present study demonstrated that greater involvement of patients with SLE in SDM contributes to greater trust in their attending rheumatologists and general physicians. Thus, the present study demonstrates the theoretical importance of SDM in the management of SLE in terms of long-term physician-patient relationship.

Previous studies on the effectiveness of SDM, particularly in the management of rheumatic diseases, have focused on the physician-patient relationship. Previous studies conducted in other clinical fields have revealed that better SDM may be associated with reduced decisional conflict, [22] increased patient knowledge, [23] selection of treatment choices consistent with their values, [23] better general health, and improved symptoms. [5] In contrast, a systematic review of long-term conditions revealed no improvement in the physician-patient relationship following interventions to support SDM. [24] A previous study conducted in the UK revealed that better SDM resulted in greater trust in the attending rheumatologist among patients with lupus nephritis; however, confounding factors, such as income and disease activity, were not adjusted for in this study. [10] A previous study conducted in the US revealed a relationship between patient-participatory decision-making and trust in physicians among patients with SLE and RA; however, this study focused on trust in general physicians, and multivariate factors were not adjusted for in the analyses. [25] Similarly, an association was observed between low SDM and low trust in attending rheumatologists among patients with RA confounded by income. [26]

These observed associations were based on cross-sectional studies; thus, reverse causation may be possible. The present study demonstrated, for the first time, that greater involvement of patients with SLE in SDM contributes to the subsequent formation of trust in their attending rheumatologists and general physicians.

The present study has several clinical implications for clinicians and policymakers. First, an improvement in trust in rheumatologists through deep involvement in SDM may improve the behavioral and health outcomes. Previous studies have revealed that greater trust in attending rheumatologists is associated with better medication adherence among patients with SLE. [7] However, further studies must be conducted to determine whether the improvement in trust in attending rheumatologists following SDM affects the future quality of life and disease activity. Second, the involvement of patients with SLE in SDM is modifiable. For instance, patients become better aware of concerns regarding disease activity and treatment effectiveness or possible side effects as they gather online health information. This enables them to express their concerns to their rheumatologists, leading to the next treatment being planned in a more collaborative manner. [4] Therefore, rheumatologists should communicate with patients to ensure that the patients are comfortable with sharing their concerns. Indeed, even disease activity is recognized differently by physicians and patients with SLE. [27] Third, the long-term improvement in trust in general physicians following their deep involvement in SDM suggests that interaction with a single physician may influence patients’ attitudes toward general physicians. Trust in general physicians is influenced by social cues, such as media portrayals and second-hand experiences, including conversations with others. [28,29] However, the findings of the present study reinforce the theory that trust in general physicians is also dependent on the primary experiences of the patients with their physicians. [29] Therefore, encouraging individual rheumatologists to practice SDM in accordance with the guidelines and recommendations may aid in improving trust in rheumatologists in general. Fourth, policymakers must incorporate the importance of SDM into guidelines and statements on the management of SLE and lupus nephritis more proactively as it increases rheumatologists’ awareness of SDM. The Joint Recommendations on the Management of Lupus Nephritis by EULAR, the European Society of Nephrology, and the European Society for Dialysis and Transplantation, [30] as well as statements on the management of SLE by EULAR or the Asia Pacific League of Associations for Rheumatology, [2,31] have recognized SDM as an overarching principle. However, SDM has not been mentioned in the American College of Rheumatology Guidelines for the Management of Lupus Nephritis or the Japanese Guidelines for the Management of SLE. [32,33]

The present study has several strengths. First, the present study demonstrated an association between greater involvement in SDM and greater trust at 1 year using two different instruments. The use of two instruments to measure trust in attending physicians ensured the robustness of this association. Second, the present study demonstrated the relationship between SDM and trust in physicians across multiple centers in Japan and adjusted for socioeconomic factors, such as education and income, thereby ensuring high internal validity and generalizability.

Nevertheless, this study had several limitations. First, the causal relationship between SDM and trust in physicians was not as strong as that in interventional studies, as this was an observational study. Second, all facilities included in the present study were university medical centers; thus, rheumatologists and patients with SLE may differ from their counterparts in clinics and general hospitals. However, patients with SLE generally visit specialized medical centers such as university hospitals in Japan.

In summary, the present study demonstrated the potential role of SDM in building trust in physicians among patients with SLE, underscoring the significance of SDM in ensuring confidence in decision-making in the management of SLE, where there is uncertainty regarding treatment efficacy and multiple treatment options.

## Data Availability

All data produced in the present study are available upon reasonable request to the authors.

## ACKNOWLEDGMENTS

We thank all colleagues in our laboratories and the members of the facilities participating in the TRUMP2-SLE project for patient recruitment, data collection, suggestions, and meaningful discussions. We also thank Editage (www.editage.com) for the English language editing. Part of this study was presented at the 67th Annual Scientific Meeting of the Japan College of Rheumatology and published as a supplementary abstract (Yoshimi R, et al. *Mod Rheumatol*. 2023; 33[Supplement]: S124).

## REFERENCES

1 Morrison T, Foster E, Dougherty J, et al. Shared decision making in rheumatology: A scoping review. Semin Arthritis Rheum 2022;56:152041. doi:10.1016/j.semarthrit.2022.152041.

2 Fanouriakis A, Kostopoulou M, Alunno A, et al. 2019 update of the EULAR recommendations for the management of systemic lupus erythematosus. Ann Rheum Dis 2019;78:736–45. doi:10.1136/annrheumdis-2019-215089.

3 Fanouriakis A, Kostopoulou M, Andersen J, et al. EULAR recommendations for the management of systemic lupus erythematosus: 2023 update. Ann Rheum Dis 2024;83 :15–29. doi:10.1136/ard-2023-224762.

4 Ichikawa T, Kishida D, Shimojima Y, et al. Impact of online health information-seeking behavior on shared decision-making in patients with systemic lupus erythematosus: The TRUMP2-SLE project. Lupus 2023;:9612033231200104;32:1258–66. doi:10.1177/09612033231200104.

5 Shay LA, Lafata JE. Where is the evidence? A systematic review of shared decision making and patient outcomes. Med Decis Making 2015;35:114–31. doi:10.1177/0272989X14551638.

6 Toupin-April K, Barton J, Fraenkel L, et al. Toward the development of a core set of outcome domains to assess shared decision-making interventions in rheumatology: Results from an OMERACT Delphi survey and consensus meeting. J Rheumatol 2017;44:1544–50. doi:10.3899/jrheum.161241.

7 Kurita N, Oguro N, Miyawaki Y, et al. Trust in the attending rheumatologist, health-related hope and medication adherence among Japanese systemic lupus erythematosus patients. Rheumatol (Oxf Engl) 2023;62:2147–53. doi:10.1093/rheumatology/keac565.

8 Mosley-Williams A, Lumley MA, Gillis M, et al. Barriers to treatment adherence among African American and white women with systemic lupus erythematosus. Arthritis Rheum 2002;47:630–8. doi:10.1002/art.10790.

9 Case S, Sinnette C, Phillip C, et al. Patient experiences and strategies for coping with SLE: A qualitative study. Lupus 2021;30:1405–14. doi:10.1177/09612033211016097.

10 Georgopoulou S, Nel L, Sangle SR, et al. Physician-patient interaction and medication adherence in lupus nephritis. Lupus 2020;29:1168–78. doi:10.1177/0961203320935977.

11 Entwistle V. Trust and shared decision-making: An emerging research agenda. Health Expect 2004;7:271–3. doi:10.1111/j.1369-7625.2004.00304.x.

12 Goto Y, Miura H, Son D, et al. Psychometric evaluation of the Japanese 9-item shared decision-making questionnaire and its association with decision conflict and patient factors in Japanese primary care. JMA 2020;3:208–15. doi:10.31662/jmaj.2019-0069.

13 Kriston L, Scholl I, Hölzel L, et al. The 9-item Shared Decision Making Questionnaire (SDM-Q-9). Development and psychometric properties in a primary care sample. Patient Educ Couns. patient ed.uc Couns 2010;80:94–9 2010;80:94–9. doi:10.1016/j.pec.2009.09.034.

14 Décary S, Toupin-April K, Légaré F, et al. Five golden rings to measure patient-centered care in rheumatology. Arthritis Care Res (Hoboken) 2020;72;Suppl 10:686–702. doi:10.1002/acr.24244.

15 Thom DH, Ribisl KM, Stewart AL, et al. Further validation and reliability testing of the Trust in Physician Scale. The Stanford trust study physicians. Med Care 1999;37:510–7. doi:10.1097/00005650-199905000-00010.

16 Suzuki R, Yajima N, Sakurai K, et al. Association of patients’ past misdiagnosis experiences with trust in their current physician among Japanese adults. J Gen Intern Med 2022;37:1115–21. doi:10.1007/s11606-021-06950-y.

17 Oguro N, Suzuki R, Yajima N, et al. The impact that family members’ health care experiences have on patients’ trust in physicians. BMC Health Serv Res 2021;21:1122. doi:10.1186/s12913-021-07172-y.

18 Dugan E, Trachtenberg F, Hall MA. Development of abbreviated measures to assess patient trust in a physician, a health insurer, and the medical profession. BMC Health Serv Res 2005;5:64. doi:10.1186/1472-6963-5-64.

19 Oguro N, Yajima N, Miyawaki Y, et al. Effect of communicative and critical health literacy on trust in physicians among patients with systemic lupus erythematosus (SLE): The TRUMP2-SLE project. J Rheumatol 2023;50:649–55. doi:10.3899/jrheum.220678.

20 Oguro N, Yajima N, Ishikawa Y, et al. Effect of attending rheumatologists’ big 5 personality traits on patient trust in patients with systemic lupus erythematosus: The TRUMP2-SLE project. J Rheumatol 2023. doi:10.3899/jrheum.2023-0603.

21 Austin PC, White IR, Lee DS, et al. Missing data in clinical research: A tutorial on multiple imputation. Can J Cardiol 2021;37:1322–31. doi:10.1016/j.cjca.2020.11.010.

22 McAlpine K, Lewis KB, Trevena LJ, et al. What is the effectiveness of patient decision Aids for cancer-related decisions? A systematic review subanalysis. JCO Clin Cancer Inform 2018;2:1–13. doi:10.1200/CCI.17.00148.

23 Stacey D, Légaré F, Lewis K, et al. Decision aids for people facing health treatment or screening decisions. Cochrane Database Syst Rev 2017;4:CD001431. doi:10.1002/14651858.CD001431.pub5.

24 Mathijssen EGE, van den Bemt BJF, van den Hoogen FHJ, et al. Interventions to support shared decision making for medication therapy in long term conditions: A systematic review. Patient Educ Couns. patient ed.uc Couns 2020;103:254–65 2020;103:254–65. doi:10.1016/j.pec.2019.08.034.

25 Berrios-Rivera JP, Street RL Jr, Garcia Popa-Lisseanu MG, et al. Trust in physicians and elements of the medical interaction in patients with rheumatoid arthritis and systemic lupus erythematosus. Arthritis Rheum 2006;55:385–93. doi:10.1002/art.21988.

26 Barton JL, Trupin L, Tonner C, et al. English language proficiency, health literacy, and trust in physician are associated with shared decision making in rheumatoid arthritis. J Rheumatol 2014;41:1290–7. doi:10.3899/jrheum.131350.

27 Yen JC, Neville C, Fortin PR. Discordance between patients and their physicians in the assessment of lupus disease activity: Relevance for clinical trials. Lupus 1999;8:660–70. doi:10.1191/096120399680411362.

28 Boulware LE, Cooper LA, Ratner LE, et al. Race and trust in the health care system. Public Health Rep 2003;118:358–65. doi:10.1093/phr/118.4.358.

29 Hall MA, Camacho F, Dugan E, et al. Trust in the medical profession: Conceptual and measurement issues. Health Serv Res 2002;37:1419–39. doi:10.1111/1475-6773.01070.

30 Fanouriakis A, Kostopoulou M, Cheema K, et al. 2019 Update of the Joint European League Against Rheumatism and European Renal Association-European Dialysis and Transplant Association (EULAR/ERA-EDTA) recommendations for the management of lupus nephritis. Ann Rheum Dis 2020;79:713–23. doi:10.1136/annrheumdis-2020-216924.

31 Mok CC, Hamijoyo L, Kasitanon N, et al. The Asia Pacific League of Associations for Rheumatology consensus statements on the management of systemic lupus erythematosus. The Lancet Rheumatol 2021;3:e517–31. doi:10.1016/S2665-9913(21)00009-6.

32 Hahn BH, McMahon MA, Wilkinson A, et al. American College of Rheumatology guidelines for screening, treatment, and management of lupus nephritis. Arthritis Care Res 2012;64:797–808. doi:10.1002/acr.21664.

33 Japan College of Rheumatology. Guideline for the management of systemic lupus erythematosus 2019 (in Japanese). Tokyo: : Nanzando 2019.

